# Even A Worm Will Turn: Appendicitis Malpractice Litigation Since 2020

**DOI:** 10.1101/2024.07.11.24310287

**Authors:** Rahma Menshawey, Esraa Menshawey

## Abstract

**Background:** Appendicitis is the inflammation of the vermiform appendix, and it is the most common abdominal surgical emergency in the world. Its diagnosis, however, has many pitfalls including lack of a pathognomonic sign or symptoms, and the low predictive value of laboratory testing. Appendicitis is a leading cause of malpractice concerns.

**Methods:** Using Google Case Law, we used the search terms “appendicitis”, and “malpractice” to identify appendicitis litigation cases. We included cases published since 2020. We included any case where a confirmed diagnosis of appendicitis was made. We included cases filed for malpractice due to complications of appendicitis and its treatment. Outcomes of interest included: the state the case was published, the defendants, the date of when the patient first complained of abdominal pain to when they had had an appendectomy/treatment, diagnostics and consultations, medical and legal issues, and final verdict/opinion/decisions on the cases.

**Results:** A total of 44 cases were identified, which were screened for inclusion. A total of 14 cases met the inclusion criteria and were analyzed. Most cases did not present in an atypical way, the majority of patients presented with clear statements of abdominal pain of varying severity. The majority of the defendants were MD’s and hospitals. The average time from symptom to diagnosis was 2.4 ±2.1 days, while the longest time for diagnosis was 7 days. The leading medico-legal issues were failure to diagnose and delayed diagnosis, while among the cases, 35.7% had outcomes in favor of the plaintiff.

**Conclusions:** Appendicitis remains an area of high risk of litigation. Malpractice suits are often due to failure to diagnose and failure to treat, but there maybe proactive measures to address the modern pitfalls to promote a decreased litigation risk and patient safety

## Introduction

“Out—out are the lights—out all! And, over each quivering form, The curtain, a funeral pall, Comes down with the rush of a storm, While the angels, all pallid and wan, Uprising, unveiling, affirm That the play is the tragedy, “Man,” And its hero, the Conqueror Worm.”

Edgar Allan Poe, The Conqueror Worm

Appendicitis is a common surgical emergency characterized by the inflammation of the vermiform appendix. It represents a significant healthcare burden globally and necessitates prompt diagnosis and treatment to prevent potentially life-threatening complications^1,2^. However, the diagnosis of appendicitis has some pitfalls, and the condition has become a leading cause of medical malpractice concerns^3^.

The accurate and timely diagnosis of appendicitis poses challenges to healthcare professionals due to several factors^4^. Firstly, there is an absence of pathognomonic signs or symptoms, and this makes it difficult to rely solely on clinical presentation, and atypical presentation can further convolute these cases^3^. Patients may present with a range of symptoms, including abdominal pain, nausea, vomiting, and fever, which can overlap with other gastrointestinal, obstetric, and genitourinary conditions^1,5–8^. Secondly, laboratory tests, such as white blood cell count and C-reactive protein levels, while useful as supportive evidence, may have a poor predictive value for appendicitis, and so they cannot be relied on alone to rule in appendicitis^9^. CT scan can be an excellent tool to diagnose appendicitis, but may not always be available, is costly, and exposes the patient to radiation^10–12^. A diagnostic laparotomy can also be used to identify and immediately treat appendicitis and its complications^11^. However, this poses the dilemma of balancing the risks of a potentially unnecessary surgery with the potential for delayed or missed diagnoses.

As a result of these pitfalls, appendicitis is a major cause of medical malpractice claims due to issues such as misdiagnosis, delayed diagnosis, and complications of the condition and its treatment^13^. Understanding the scope, as well as the characteristics of appendicitis litigation is crucial for identifying areas of concern and improving patient outcomes and safety. Therefore, the purpose of this study is to analyze medical malpractice cases involving appendicitis published since 2020. We seek to identify trends and characterize the appendicitis litigation findings and outcomes which form these cases.

## Methods

Using Google Scholar Case Law, we searched for “appendicitis”, and “malpractice” on 2024-07-04. We included all cases in our search posted since 2020 to 07-2024. A total of 44 cases were identified. Each case was manually screened by the authors and analyzed. A narrative summary of the cases can be found in a supplementary file. Cases that did not list medically relevant defendants were excluded. We included cases where the condition of appendicitis was the primary issue in the litigation case (see figure 1). We also included cases that made claims on the complications of appendicitis and its treatment. All cases analyzed where in the English language and filed in the United States of America. We excluded any case updates (only used the latest published case filing for the analysis). We gathered data on the patient’s presentation date (to the ER or otherwise), the date of appendicitis treatment or appendectomy, explicitly mentioned symptoms, diagnostic tests (X-ray, ultrasound, CT scan), medicolegal issues, and the final verdict/opinion/orders of each case. (See table 1) Legal verdicts and medico-legal issues are listed in table 2 (See table 2). We also conducted a review of the literature on appendicitis malpractice.

**Figure 1.**
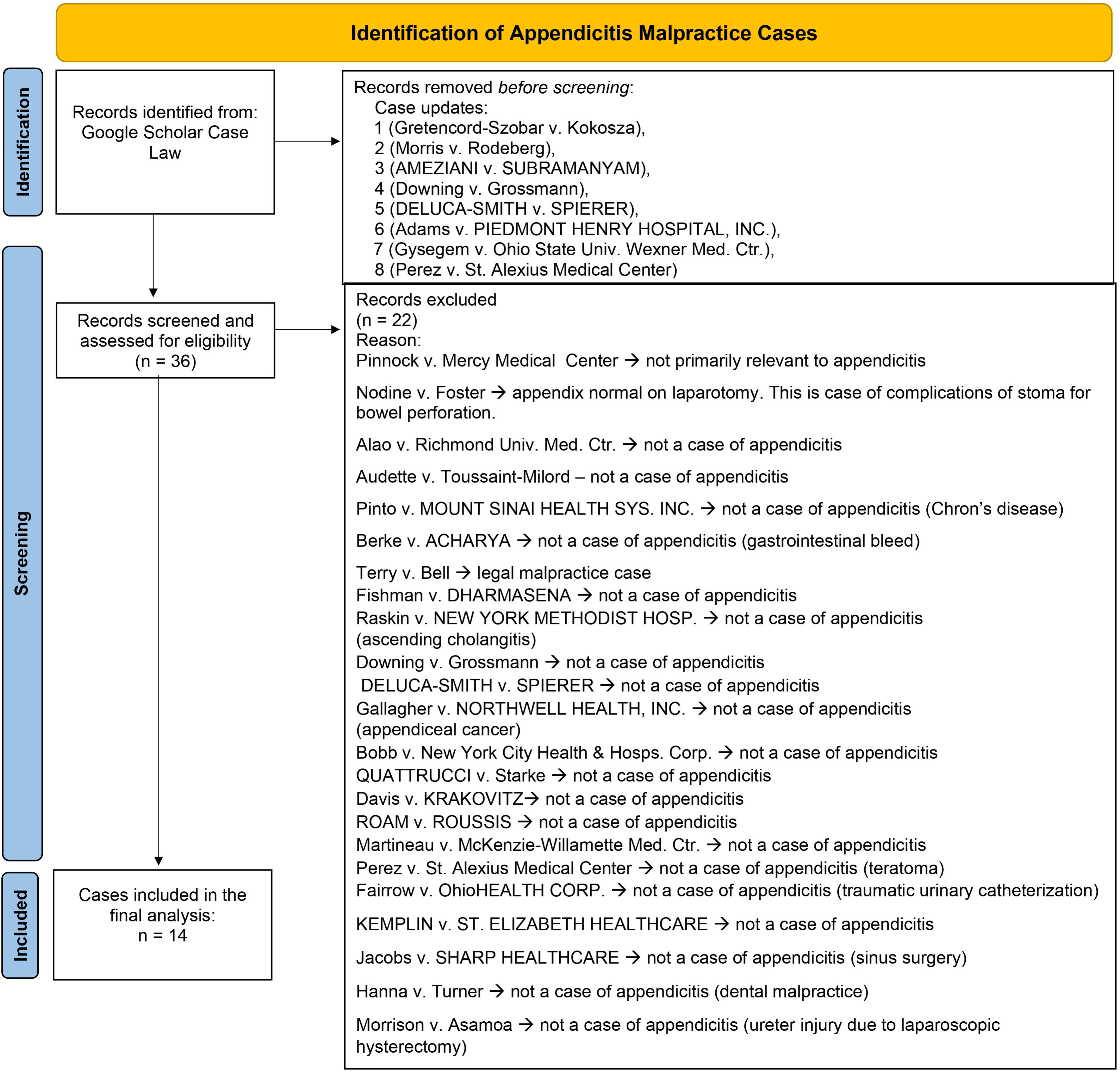
Flow chart depicting the methodology for finding cases using the Google Case Law database. accessed 2024-07-04. A total of 44 cases were identified by the search engine using the words “appendicitis” and “malpractice”. The final analysis was conducted on 14 cases which met the inclusion criteria. Reasons for exclusion of other cases are found within the chart.

**TABLE 1:** Case details and outcomes of interest. P.C: professional corporation, (Cases in green background denote a verdict/decision.order in favor of the defendant, while red denotes a verdict in favor of the plaintiff). Day of treatment includes the day either when appendicitis was diagnose, or when a CT scan was performed which showed signed of appendicitis, or when the appendectomy took place. Symptoms were extracted from the case filing as they were exactly mentioned.

| Case | STATE | Defendants | Notable symptoms | CT obtained? |
| --- | --- | --- | --- | --- |
| Gretendcord-Szobar v. Kokoszka | Illinois | Surgical consultant<br>Surgical associates | Severe Abdominal Pain<br>On examination: Lower quadrant abdominal pain | YES – one CT showed normal appendix<br>A later CT revealed an enlarged appendix |
| DE DURAN v. FOREST HILLS HOSP. | New York | Hospital<br>Health systems<br>RN<br>RN<br>MD | nausea, pain and vomiting<br>pain 7/10 | No |
| PANTALEON v. Garber | New York | MD – resident physician<br>MD<br>MEDICAL CENTER | Abdominal pain<br>No Mcburney point tenderness, rebound, guarding, or fever was noted. | No |
| AMEZIANI v. SUBRAMANYAM | New York | MD<br>MD | Sharp intense abdominal pains | No |
| Hensley v. FROEDTERT SOUTH, INC. | Wisconsin | HOSPITAL<br>HOSPITAL<br>MD<br>DO | lower abdominal and back pain<br>bloating, nausea,<br>diarrhea, vomiting, constipation, recurring fever. | No |
| Morris v. Rodeberg (pediatric) | North Carolina | MD,<br>Hospital | Right sided abdominal pain | No |
| MYKLYN v. CONEY ISLAND HOSPITAL | New York | Hospital corporation<br>Hospital | Abdominal pain, Nausea, Vomiting, Mild fever<br>Back ache | No |
| Richter v. MENOCAL | New York | MD<br>HOSPITAL | No mention | No mention |
| GUYETT v. KALEIDA<br>HEALTH | New York | MD<br>Hospital<br>Health network | No mention | No mention |
| Adams v. PIEDMONT<br>HENRY HOSPITAL,<br>INC. | Georgia | Hospital Inc.<br>Healthcare Inc.<br>MD<br>Medical P.C<br>Surgery P.C | Abdominal pain | No Mention |
| Lewis v.<br>WIGNAKUMAR | Kentucky | MD | Severe pain in the stomach/abdomen | Yes |
| Gysegem v. Ohio State<br>Univ. Wexner Med. Ctr. | Ohio | University medical center | Pain | Yes – CT scan showed “extraluminal collection containing an air-fluid level adjacent to the appendix with an appendicolith in this region... consistent with a contained fluid collection secondary to perforated appendicitis”. |
| Hill v. Springfield Hosp. | Vermont | Hospital<br>Emergency services | No mention | No mention |
| Vilas v. Love | Tennessee | MD | Pain | No mention |

**TABLE 2:**
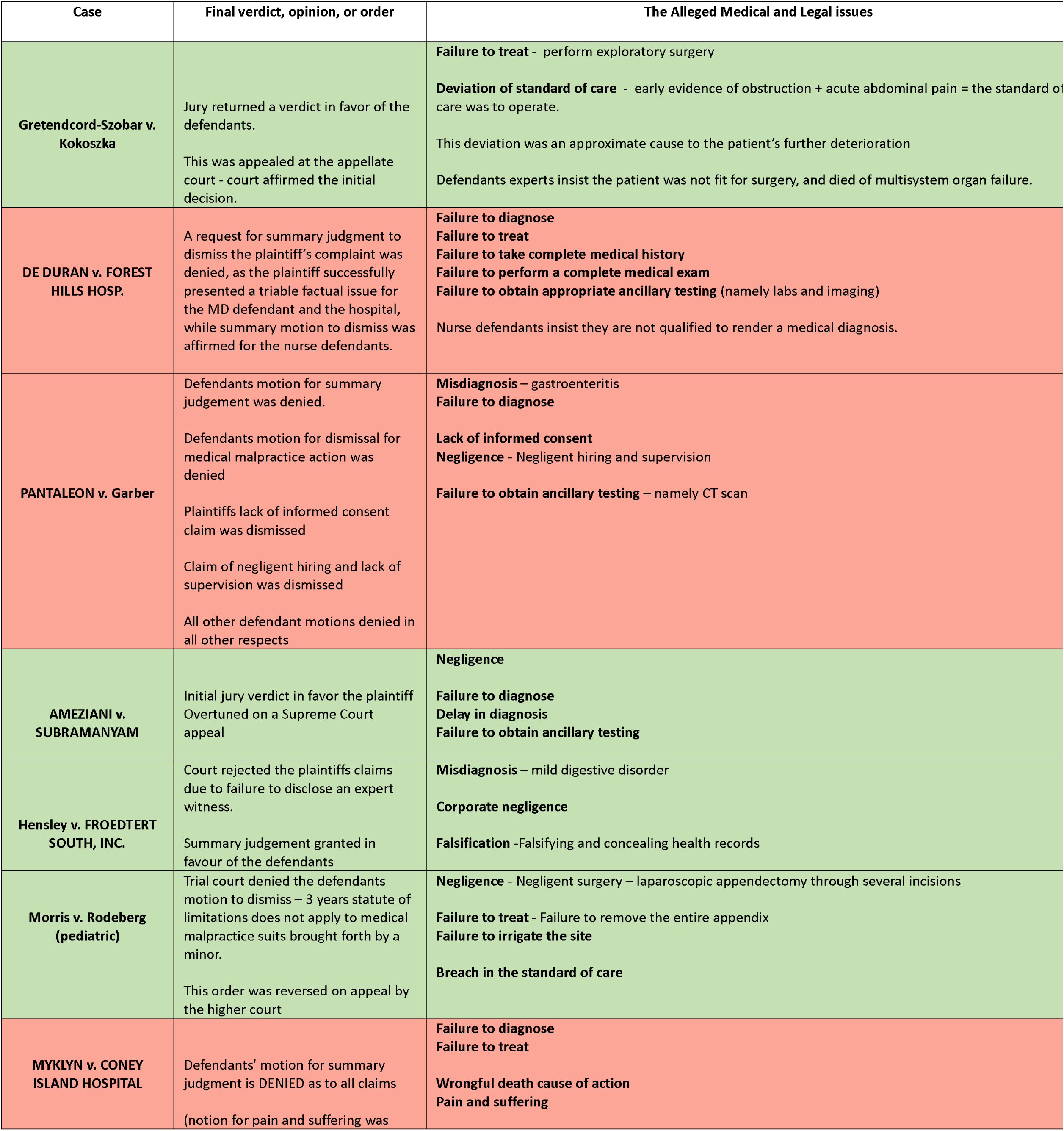
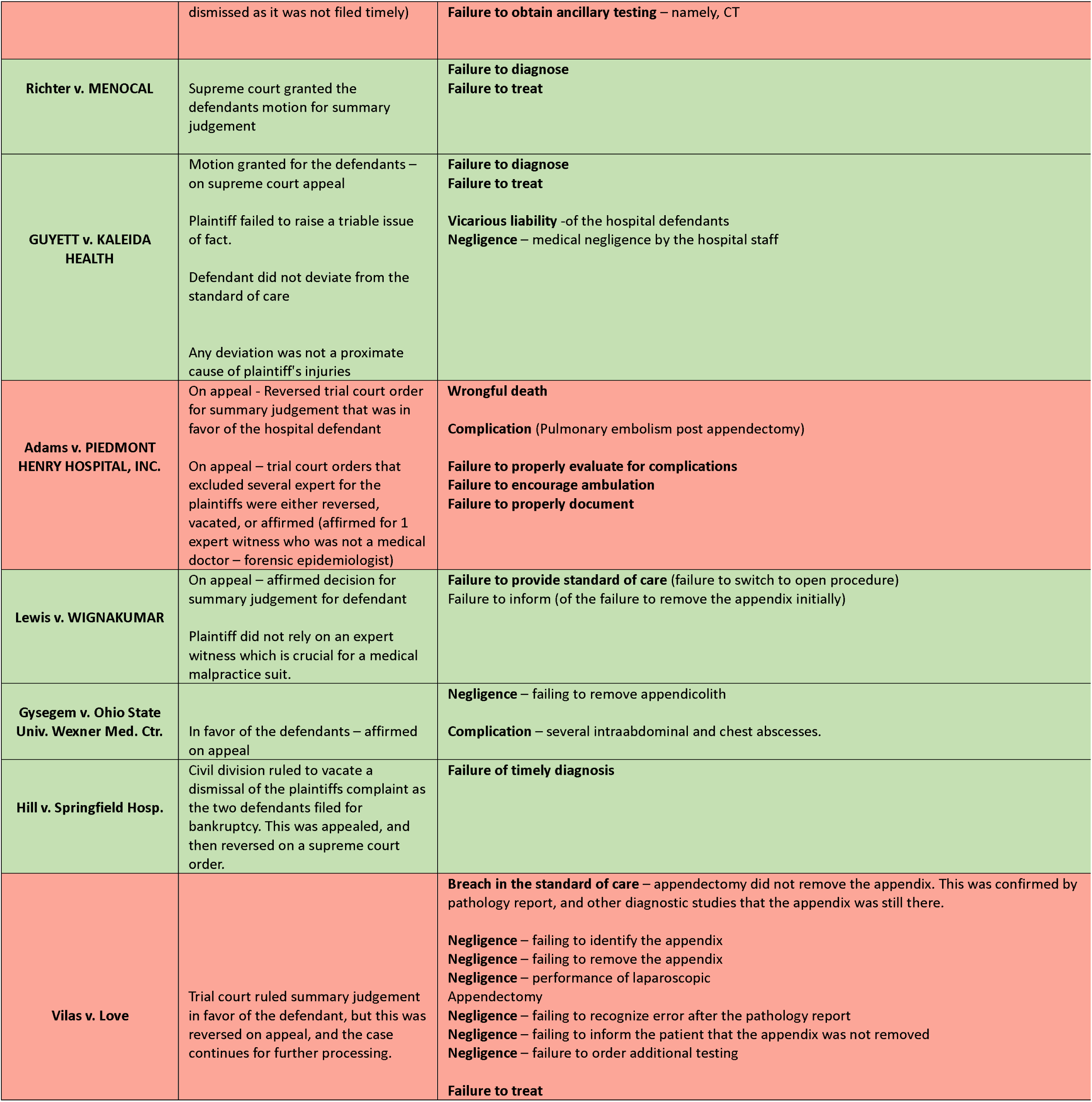
Medico-legal outcomes of the case, as well as the final verdict, opinion in order identified in the latest case publication.

## Results

A total of 14 cases met our inclusion criteria. Cases were filed across 9 different states (see figure 2), and 43% of cases were filed in the state of New York (n = 6) (see figure 2).

**Figure 2.**
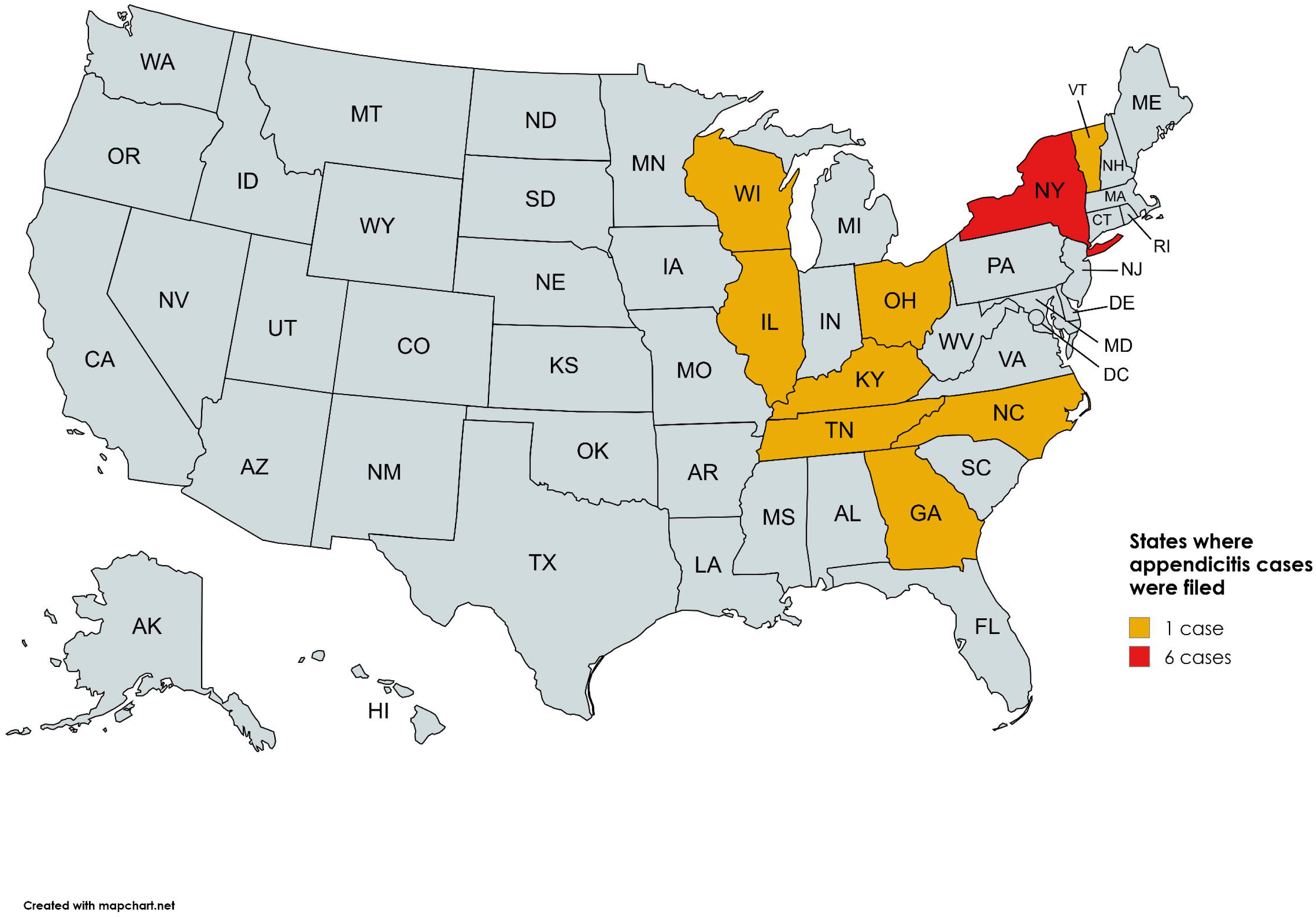
Map showing the which states the cases originated from. The majority of appendicitis malpractice cases were overwhelmingly filed in the state of New York.

Several defendants were listed in the cases. There was a total of 35 defendants across all 14 cases, which comes to an average of 2.5 defendants per case. The majority of defendants were medical doctors (40%, n = 14), one of which was a second-year surgery resident. Two registered nurses were listed as defendants. Hospitals were there second most commonly cited defendants (22.86%, n=8). The rest of the defendants included emergency service, health networks, health systems, health care incorporation’s, medical centers, medical and surgical professional corporations, and surgery associates.

Among the cases, 10 cases mentioned an exact date for when the patient was presented for care, and when they were diagnosed or treated. The average time from those two points was 2.4 ±2.1 days. The longest time to diagnosis was 7 days, while in the majority of cases, 60%, were diagnosed and treated within a day of presentation (n=6), among the cases which listed exacted dates.

Several notable symptoms were noted, including (as they are mentioned); severe abdominal pain, right lower quadrant pain, sharp intense abdominal pain, back pain, back ache, bloating, nausea, and vomiting. Among the cases that detailed the patient’s symptom (n=11), all of them, 100%, mentioned the symptoms of pain on presentation, and so we can surmise that atypical presenting appendicitis is not the trigger for malpractice claims. Only one case, which also presented with pain, made a mention of lack of McBurney point tenderness which was the basis of a misdiagnosis.

Diagnostic imaging data was also extracted from the cases including Xray’s, abdominal ultrasound, and CT scans. Only 3 cases explicitly mentioned the use of a CT scan, and all 3 of those cases ended with a verdict in favor of the defendants. The lack of obtaining a CT was occasionally mentioned in the malpractice claims and a point of faulting the defendant by the plaintiff’s expert witness due to the excellent sensitivity of the CT scan in diagnosing appendicitis, mixed with the reliance only on labs or clinical findings, which maybe poorly predictive for appendicitis.

Table 2 lists the opinion or verdict or decision outcomes of each case. Among the cases, 35.7% had outcomes in favor of the plaintiff (n=5). Appendicitis cases pose a tough litigation and can go far, as several of the cases continued to appeal trial court decisions, and several also involved supreme court decisions, and others appealed those decisions. Cases often turned into a battle of expert opinions, and argument ensued over several points including the findings on diagnostic imaging like CT scan, the reliance of the defendant on only labs and the lack of fever, the failure to act on increased white blood count, the decision to rely on oral temperature rather than rectal temperature, infrequent vital sign examination, the decision to not immediately operate, the decision to do partial or complete irrigation during surgery, and in the management of complications.

Several of the cases were dismissed or motion granted to the defendant because the plaintiff patient believed they can rely solely on the facts of the case, or a preponderance of evidence, or if their care was against direct hospital policy or guidelines (such as a hospital policy to proceed with surgery for all patients with abdominal pain and fever, rather than medical therapy). It can be stated with confidence that the failure of the plaintiff to resort to a medical expert witness will doom their case. Expert witnesses are extremely critical in appendicitis cases, where some cases even greatly disputed the qualifications and credentials of them, and tremendous argument ensured over the aforementioned details. Additionally, appendicitis cases often require several expert witnesses on both sides and these include emergency physicians, pediatricians, infectious disease experts, surgeons, physicians involved in specialties relating to complications of treatment (such as a cardiologist in the treatment of pulmonary embolism as a post appendectomy complication), forensic epidemiologists, and more.

Plaintiffs faltered in their cases due to other reasons including the statute of limitations, and this is especially notable in the only case involving minor who tried to bring forth a medical malpractice suit (see Morris v. Rodeberg). Suing as a result of a complication was another challenge especially if it is a potentially common complication such as an intra-abdominal abscess, where defendants and their expert claim that this is possible due to the difficulty that can arise in removing an infected nidus which can result in an abscess and their reoccurrence. Two of the examined cases made a claim for the complications post op, several abscess reoccurrences (due to a retained appendicolith) which was won for the defendants, and the other was a medical malpractice and wrongful death claim due to a pulmonary embolism post-appendectomy and abysmal pre-op and post-op preventative care, which was won for the plaintiff on appeal.

There are several alleged medico-legal issues in the examined cases, which include; failure to treat, deviation of the standard of care, failure to diagnose, delay in the diagnosis, failure to obtain a complete medical history and physical examination, failure to obtain additional diagnostic testing (namely labs, and a CT scan), misdiagnosis, lack of informed consent, negligence (negligent hiring, negligent supervision, corporate negligence), falsification and or concealment of records, wrongful death, and pain and suffering.

## Discussion

Our analysis of appendicitis litigation cases highlights the complex interplay of medico-legal challenges and underscoring the pivotal factors and findings that shape them. In this study, we examined the modern litigation surrounding appendicitis cases, aiming to characterize the issue. Our analysis focused on a total of 14 cases that met our inclusion criteria, filed across 9 different states. Among the cases, we identified multiple defendants, including medical doctors, registered nurses, and hospitals, among others. Furthermore, the average time of 2.4 ± 2.1 days from presentation to diagnosis. Severe abdominal pain and other associated symptoms were consistently reported by the majority of patients. Additionally, the verdicts or decisions varied, with 35.7% of cases favoring the plaintiff. Appendicitis cases can go far, with the majority of cases seeking appeal, supreme court verdict, and even supreme court appeal. These findings underscore the complexity of appendicitis litigation.

A literature review reveals interesting findings. Fear of malpractice is a recognized concern that may drive unnecessary and defensive practices. Emergency care^14^, as well as radiology^15^, is moderately affected by appendicitis litigation among other conditions^16^. Appendicitis consistently ranks among the top diagnosis-related malpractice claims in the United States. Misdiagnosis of gastroenteritis appears to be a common pitfall^17^.

Appendicitis litigation is a critical issue among the pediatric population^18–20^. One study found that the failure or delays in diagnosing appendicitis were the most common causes of malpractice lawsuits, and resulted in the largest payments to the plaintiff^21^. Likewise, the leading issue that triggered the malpractice suit in our examined cases was also failure or delayed diagnosis (rather than misdiagnosis). Adult patients may make up the majority of defendants in the cases we examined for several reasons including increased awareness and information that can encourage them to take suit (even to a detriment point where they do not seek out an expert witness, relying only on the facts of the case, which can doom the case outcome), and their increased understanding of their case and symptoms causing them to be more proactive in seeking legal recourse (which is a common finding in the cases we analyzed where several of adult plaintiffs filed claims the following day on recognizing the alleged malpractice), as well as the prolonged long term impact of a mismanaged case which can also inspire legal action (such as in the case of Gysegem v. Ohio State Univ. Wexner Med. Ctr. where the plaintiff continuously suffered from recurrent abscess which resulted in their liver fully adhering to the anterior abdominal wall, and abscess breaching the diaphragm and chest cavity to encompass the lung).

CT scan is a highly sensitive diagnostic modality to diagnose appendicitis but has complications like radiation exposure which should be minimized or avoided in children and pregnant women. Additionally, some studies point to increased sensitivity with increased radiation doses^10–12^. In our study, the 3 cases that specifically mentioned the use of a CT all ended with a verdict favorable for the defendant. In the cases where it was not used, this point was consistently brought up by the plaintiff’s expert witness as a breach in the standard of care. It may seem that the use of a CT scan may provide a “good” defensible position, but this tricky territory, and would need to be confirmed by further studies. Additionally, false positives and false negative results can occur with the use of CT (one of the excluded cases involved a misdiagnosis of appendicitis, resultant appendectomy, and further complications for a patient with Crohn’s disease).

Sosner et al suggest the following considerations:

> a focused CT may not always capture the appendix
>
> an appendix may not be appreciated in a larger person
>
> the appendix must be examined carefully and fully from the origin to the distal tip
>
> caliber alone is not an indicator for appendicitis, look for haziness of regional fat, peri-appendiceal fluid, and perforation and rupture for evidence of appendicitis
>
> check unusual locations for the appendix such as in a hernia sac^22^.

A Canadian study also suggests the following during ultrasound to improve the visualization of the appendix; 1) identify the maximum point of tenderness to locate the quadrant for assessment, 2) use the ileocecal valve as a marker as the appendix should be inferior and medial, 3) push the cecum medially with probe to visualize the appendix if suspecting a retrocecal position, and 4) using your other hand to provide counterpressure and redistribute bowel gases^23^.

In a review of 234 appendicitis malpractice cases, from 1983 to 2012, the majority of cases cited a delay in diagnosis, followed by intraoperative negligence^24^. A delayed diagnosis led to rupture, death, and fertility issues. We also observed cases where there was intraoperative negligence including failure to completely remove the appendix, failure to remove an appendicolith, failure to irrigate the area, and removal of inflamed epiploic appendage instead of appendix. In one case, Vilas v. Love, it was only discovered on the pathology report that there was no appendix in the tissue sample, and the patient later discovered their appendix was never disturbed in the previous operation. On appeal, the summary judgement granted for the defendant was reversed.

Overall, a delay in diagnosis can be due to^25^; atypical presentation, lack of a thorough physical examination including a rectal examination (one case made mention of rectal temperature preference over oral), those receiving narcotics which can mask the pain symptoms, and those diagnosed with gastroenteritis even in the absence of typical symptoms (PANTALEON v. Garber).

“Good records equals a good defence” is a wise adage, as in many cases poor documentation was another pitfall. Unclear statements, or statements that are not specific can be heavily argued. In one notable case AMEZIANI v. SUBRAMANYAM, the radiological report stated “no abnormality” with regards to the lower abdominal cavity. This seems to have resulted in some confusion, and a delay in care. The jury deliberated if a statement of “no abnormality” could have misled the treating physician as it was vague as to whether this included the appendix, and they agreed, but this was overturned only on a supreme court appeal. Agreement between the nurses notes and the physicians is also important^26^. All interactions involving the patient care should be documented, while all new changes in lab findings should be proactively and immediately addressed or investigated. Meanwhile, it is important to maintain a broad differential when it comes to abdominal pain, while one should be wary with the use of pain medication as it can mask important signs and symptoms or provide a false sense of reassurance^26^.

Appendicitis malpractice litigation is also a global problem. In Germany for example, pediatric appendicitis malpractice claims analysed over a 5 year period revealed that the most common reasons for conviction were surgical technical errors, and delayed diagnosis^27^. To decrease the risk of litigation, the authors recommend the objective and professional documentation of all interactions, to accept ones limits and seek help for unfamiliar cases, and to maintain transparent communication with parents and family members^27^.

Meanwhile in England, one study found that preoperatively, misdiagnosis and delayed diagnosis were leading, intraoperative issues were due to faulty surgical technique, while post operative claims were due to complications, inadequate follow-up and infections. damages to fertility were another claim^28^. In cases where there was a failure to take action in response to histological reports that revealed malignant changes, there was a 100% successful litigation rate^28^. Although this scenario was not seen in the cases we examined, the failure to act on pathological reports was a significant issue in one case when the report confirmed that there was no appendix in the tissue sample, although the physician argued that they did remove the appendix based on known landmarks, and a summary judgement was reversed on appeal for this case (Vilas v. Love). In France, appendicitis was among the top 10 diagnoses involved in pediatric malpractice claims, with a 40% mortality rate^29^. In Canada, analysis revealed that peer experts were critical of the care received by patients. Clinician factors included the failure to investigate with imaging, such as a CT scan^23^, which is also in agreement with our findings. Meanwhile, team factors critical In Japan, appendicitis in non-trauma cases was a leading cause of diagnostic errors^30^.

The literature frequently cites the state of New York with the most malpractice suits. Likewise, the majority of the cases in this study were filed in the state of New York. We believe this could be due to population density which could increase the load on hospitals and trigger delays in diagnosis, increased number of hospitals, increased medical insurance rates which could attract frivolous and costly lawsuits^31^, statute of limitations is extended in the state to 2 years and 6 months (an additional discovery rule applies for foreign bodies which have a statute of 1 year since their discovery, while minors have 2.5 years to file a claim after reaching adult hood^32,33^. Finally, there is no cap on the amount of damages that a plaintiff can claim, and this can lead to higher payouts and incentivise litigation. All these factors could explain why the state of New York is a litigation rich state.

Based on the modern cases we analysed we recommend the following considerations to mitigate the litigation risk:

- Keep professional detailed notes documenting all interactions including those with patient, nurses, other physicians, and members of the health care team
- Do not ignore changes in labs, especially white blood cell count
- Consider rectal examination and rectal temperatures. Do not rely solely on oral temperature to rule out a fever
- Seek a consult where necessary. Ask for help. The majority of examined cases did not seek out any additional consult, or of they did it was totally unrelated to the appendicitis condition. The lack of consultation or collaboration of opinions to diagnose appendicitis caused the majority of these cases to be swamped with expert opinions from all areas including surgeons, emergency physicians, radiologists, infectious disease experts, and even forensic epidemiologists. Do not wait till a court date to ask for a second opinion.
- If you think its gastroenteritis, but there is no vomiting and nausea, it might not be gastroenteritis
- Be careful with pain therapy. Morphine can mask symptoms and physical findings associated with appendicitis.
- Do not be hasty to rule out appendicitis in a patient with right lower quadrant pain. Even if there is no fever, even if there is no Mcburney point tenderness, and even if there is no rebound and guarding.
- Proper discharge is essential^34^. The patient must be well advised on what to do if they have pain, that appendicitis could be the cause of their pain, when to return to the ER, and when to go to their PCP for further follow-up

## Conclusion

Appendicitis remains a hot area of litigation, and a challenging one as both defendants and plaintiffs may go as far to defend their claims. While atypical presenting appendicitis may be a feared diagnostic pitfall, all of the cases examined had a patient presenting with abdominal pain. While appendicitis is a common pediatric surgical emergency, the majority of litigation is by adults where this condition is undiagnosed, during the time period we examined. A failure to diagnose, or a delayed diagnosis was a leading cause of litigation.

Limitations of studies on the case law can be delays in findings as such databases are continuously updated with new court documents. Additionally, legal documents are at times not overburdened with medical information. Nonetheless, for the majority of cases we are still able to surmise relevant dates, and other information such as the use of CT scan, and other findings, to help characterize the landscape of appendicitis litigation. Expert witnesses are a pivotal part of the process, often requiring several witnesses, whose qualifications and opinions may be challenged. The lack of acquiring a medical expert witness by the plaintiff will doom a case of medical malpractice claim.

Litigation studies can help point the direction of further supportive measures for providers and patients alike buy inspiring proactive steps, tools, guidelines, checklists and other means, to promote patient safety and optimal care and to minimize the risk of litigation. Our findings can used to inform clinical practice, enhance patient safety, and understand the factors that surround the occurrence of medical malpractice in appendicitis cases.

## Data Availability

All data produced in the present study are available upon reasonable request to the authors

## List of abbreviations

CT: Computer tomography
ER: Emergency room
PCP: Primary care physician

## Declarations

### Ethics Approval and Consent to Participate

Not applicable

### Consent for Publication

Not applicable

### Availability of Data and Material

All data is available in this manuscript and in additional supplementary files.

### Authors Contributions

RM conceived the idea. RM and EM compiled information, analysed the results, wrote the manuscript and approve its final form. All authors have participated significantly to this work to warrant inclusion as an author based on ICJME guidelines for authorship.

## Acknowledgements

Not Applicable

## References

1. Humes DJ, Simpson J. Acute appendicitis. BMJ. 2006;333(7567):530–534. doi:10.1136/bmj.38940.664363.AE

2. Constantin M, Petrescu L, Mătanie C, et al. The Vermiform Appendix and Its Pathologies. Cancers (Basel). 2023;15(15):3872. doi:10.3390/cancers15153872

3. Vissers RJ, Lennarz WB. Pitfalls in Appendicitis. Emerg Med Clin North Am. 2010;28(1):103–118. doi:10.1016/j.emc.2009.09.003

4. Harada T, Harada Y, Hiroshige J, Shimizu T. Factors associated with delayed diagnosis of appendicitis in adults: A single-center, retrospective, observational study. PLoS One. 2022;17(10):e0276454. doi:10.1371/journal.pone.0276454

5. Kumar A, Ramakrishnan T, Sahu S. Differential Diagnosis for Acute Appendicitis: Epiploic Appendagitis. Med J Armed Forces India. 2009;65(3):276–277. doi:10.1016/S0377-1237(09)80026-9

6. El Hentour K, Millet I, Pages-Bouic E, Curros-Doyon F, Molinari N, Taourel P. How to differentiate acute pelvic inflammatory disease from acute appendicitis[]? A decision tree based on CT findings. Eur Radiol. 2018;28(2):673–682. doi:10.1007/s00330-017-5032-4

7. Sasaki Y, Komatsu F, Kashima N, et al. Clinical differentiation of acute appendicitis and right colonic diverticulitis: A case-control study. World J Clin Cases. 2019;7(12):1393–1402. doi:10.12998/wjcc.v7.i12.1393

8. Téoule P, de Laffolie J, Rolle U, Reißfelder C. Acute Appendicitis in Childhood and Adulthood: An Everyday Clinical Challenge. Dtsch Arztebl Int. Published online November 6, 2020. doi:10.3238/arztebl.2020.0764

9. Yang H, Wang Y, Chung P, Chen W, Jeng L, Chen R. LABORATORY TESTS IN PATIENTS WITH ACUTE APPENDICITIS. ANZ J Surg. 2006;76(1-2):71–74. doi:10.1111/j.1445-2197.2006.03645.x

10. Haijanen J, Sippola S, Tammilehto V, et al. Diagnostic accuracy using low-dose versus standard radiation dose CT in suspected acute appendicitis: prospective cohort study. British Journal of Surgery. 2021;108(12):1483–1490. doi:10.1093/bjs/znab383

11. Storz C, Kolb M, Kim JH, et al. Impact of Radiation Dose Reduction in Abdominal Computed Tomography on Diagnostic Accuracy and Diagnostic Performance in Patients with Suspected Appendicitis. Acad Radiol. 2018;25(3):309–316. doi:10.1016/j.acra.2017.09.012

12. Rud B, Vejborg TS, Rappeport ED, Reitsma JB, Wille-Jørgensen P. Computed tomography for diagnosis of acute appendicitis in adults. Cochrane Database of Systematic Reviews. 2019;2019(11). doi:10.1002/14651858.CD009977.pub2

13. Téoule P, de Laffolie J, Rolle U, Reißfelder C. Acute Appendicitis in Childhood and Adulthood: An Everyday Clinical Challenge. Dtsch Arztebl Int. Published online November 6, 2020. doi:10.3238/arztebl.2020.0764

14. Poyorena C, Anderson A, Pollock JR, et al. A review of medical malpractice cases involving trainees in the emergency department. J Am Coll Emerg Physicians Open. 2023;4(4). doi:10.1002/emp2.13014

15. Baker SR, Shah S, Ghosh S. Radiology medical malpractice suits in gastrointestinal radiology: prevalence, causes, and outcomes. Emerg Radiol. 2015;22(2):141–145. doi:10.1007/s10140-014-1268-3

16. Menshawey R, Menshawey E. Knot Guilty? An Examination of Testicular Torsion Litigation Trends from 2014 to 2022. medRxiv. Published online January 1, 2024:2024.06.30.24309735. doi:10.1101/2024.06.30.24309735

17. Ferguson B, Geralds J, Petrey J, Huecker M. Malpractice in Emergency Medicine—A Review of Risk and Mitigation Practices for the Emergency Medicine Provider. J Emerg Med. 2018;55(5):659–665. doi:10.1016/j.jemermed.2018.06.035

18. Edwards BL, Dorfman D. High-risk Pediatric Emergencies. Emerg Med Clin North Am. 2020;38(2):383–400. doi:10.1016/j.emc.2020.01.004

19. Weinstock MB, Jolliff H. High-Risk Medicolegal Conditions in Pediatric Emergency Medicine. Emerg Med Clin North Am. 2021;39(3):479–491. doi:10.1016/j.emc.2021.04.001

20. Glerum KM, Selbst SM, Parikh PD, Zonfrillo MR. Pediatric Malpractice Claims in the Emergency Department and Urgent Care Settings From 2001 to 2015. Pediatr Emerg Care. 2021;37(7):e376–e379. doi:10.1097/PEC.0000000000001602

21. Sullins VF, Rouch JD, Lee SL. Malpractice in Cases of Pediatric Appendicitis. Clin Pediatr (Phila). 2017;56(3):226–230. doi:10.1177/0009922816656621

22. Sosner E, Patlas MN, Chernyak V, Dachman AH, Katz DS. Missed Acute Appendicitis on Multidetector Computed Tomography and Magnetic Resonance Imaging: Legal Ramifications, Challenges, and Avoidance Strategies. Curr Probl Diagn Radiol. 2017;46(5):360–364. doi:10.1067/j.cpradiol.2017.03.003

23. Treanor L, Drury A, Egri C, Barrett S. “Rule out appendicitis”: a Canadian emergency radiology perspective on medicolegal risks, imaging pitfalls, and strategies to improve care. Emerg Radiol. 2024;31(2):239–249. doi:10.1007/s10140-024-02214-4

24. Choudhry AJ, Anandalwar SP, Choudhry AJ, et al. Uncovering Malpractice in Appendectomies: a Review of 234 Cases. Journal of Gastrointestinal Surgery. 2013;17(10):1796–1803. doi:10.1007/s11605-013-2248-8

25. Bird S. Failure to diagnose: appendicitis. Aust Fam Physician. 2004;33(12):1025–1026.

26. Pilcher CA. When the Patient with Missed Appendicitis is Married to a Malpractice Attorney. Emergency Medicine News. 2024;46(6):18–18. doi:10.1097/01.EEM.0001024268.50619.85

27. Mahler S, Gianicolo E, Muensterer OJ. A detailed analysis of pediatric surgical malpractice claims in Germany: what is the probability of a pediatric surgeon to be accused or convicted? Langenbecks Arch Surg. 2021;406(6):2053–2057. doi:10.1007/s00423-020-02069-6

28. Mosedale T, Nepogodiev D, Fitzgerald JEF, Bhangu A. Causes and Costs of a Decade of Litigation Following Emergency Appendectomy in England. World J Surg. 2013;37(8):1851–1858. doi:10.1007/s00268-013-1907-y

29. Najaf-Zadeh A, Dubos F, Pruvost I, Bons-Letouzey C, Amalberti R, Martinot A. Epidemiology and aetiology of paediatric malpractice claims in France. Arch Dis Child. 2011;96(2):127–130. doi:10.1136/adc.2010.189209

30. Miyagami T, Watari T, Harada T, Naito T. Medical Malpractice and Diagnostic Errors in Japanese Emergency Departments. Western Journal of Emergency Medicine. 2023;24(2):340–347. doi:10.5811/westjem.2022.11.55738

31. Doesn’t liability insurance actually encourage lawsuits? The Presser Law Firm, PA. https://www.assetprotectionattorneys.com/blog/2018/january/doesnt-liability-insurance-actually-encourage-la/

32. Statute of Limitations In All 50 States MWL Law. Matthiesen, Wickert & Lehrer SC. https://www.mwl-law.com/resources/statute-limitations-50-states/

33. Schloemann M. 2024 New York Physician’s Guide to Medical Malpractice Insurance MEDPLI. MEDPLI Professional Liability Insurance. Published online March 2024. https://medpli.com/new-york-physicians-guide-to-medical-malpractice-insurance/

34. Brown-Forestiere R, Furiato A, Foresteire NP, Kashani JS, Waheed A. Acute Appendicitis: Clinical Clues and Conundrums Related to the Greatest Misses. Cureus. Published online May 11, 2020. doi:10.7759/cureus.8051

